# Genetic and environmental influences on educational disparities in adult weight change: an individual-based pooled analysis of 11 twin cohorts

**DOI:** 10.1101/2025.11.18.25340475

**Authors:** Alvaro Obeso, Gabin Drouard, Aline Jelenkovic, Emanuela Medda, Corrado Fagnani, Virgilia Toccaceli, Antti Latvala, Sari Aaltonen, Sarah E Medland, Scott D Gordon, Jooyeon Lee, Soo Ji Lee, Joohon Sung, Hyojin Pyun, Glen E Duncan, Dedra Buchwald, Juan R Ordoñana, Juan F Sánchez-Romera, Eduvigis Carrillo, Carol E Franz, William S Kremen, Robin P Corley, Brooke M Huibregtse, Patrik KE Magnusson, Ida K Karlsson, Anna K Dahl Aslan, Michael J Lyons, Meike Bartels, Lannie Ligthart, Eco JC de Geus, Margaret Gatz, David A Butler, René Pool, Anders Eriksson, Susanne Bruins, Nicholas G Martin, Dorret I Boomsma, Jaakko Kaprio, Karri Silventoinen

**Affiliations:** Helsinki Institute for Demography and Population Health, University of Helsinki, Helsinki, Finland; Department of Genetics, Physical Anthropology and Animal Physiology, Faculty of Science and Technology, University of the Basque Country (UPV/EHU), Bilbao, Spain; Institute for Molecular Medicine Finland, HiLIFE, University of Helsinki, Helsinki, Finland; Centre for Behavioural Sciences and Mental Health, Istituto Superiore di Sanità, Rome, Italy; Institute of Criminology and Legal Policy, University of Helsinki, Helsinki, Finland; QIMR Berghofer Medical Research Institute, Brisbane, Australia; Institute of Health & Environmental, Seoul National University, Seoul, Korea; Department of Epidemiology, Seoul National University School of Public Health, Seoul, Korea; Washington State Twin Registry, Washington State University - Health Sciences Spokane, Spokane, WA, USA; Washington State Twin Registry, University of Washington, Seattle, WA, USA; Department of Human Anatomy and Psychobiology, University of Murcia, Murcia, Spain; Murcia Institute for Biomedical Research (IMIB-Arrixaca), Murcia, Spain; Department of Psychiatry, University of California San Diego, La Jolla, CA, USA; Institute for Behavioral Genetics, University of Colorado, Boulder, CO, USA; Department of Psychology and Neuroscience, University of Colorado, Boulder, CO, USA; Department of Medical Epidemiology and Biostatistics, Karolinska Institutet, Stockholm, Sweden; School of Health Sciences, University of Skövde, Skövde, Sweden; Department of Psychological and Brain Sciences, Boston University, Boston, MA, USA; Department of Biological Psychology, Vrije Universiteit Amsterdam, Amsterdam, the Netherlands; Amsterdam Public Health Research Institute, Amsterdam, the Netherlands; Center for Economic and Social Research, University of Southern California, Los Angeles, CA, USA; Health and Medicine Division, The National Academies of Sciences, Engineering, and Medicine, Washington, DC, USA; cGEM, Institute of Genomics, University of Tartu, Tartu, Estonia; Complex Trait Genetics, Center for Neurogenomics and Cognitive Research, Vrije Universiteit Amsterdam, The Netherlands

**Author notes:** **Corresponding author**: Alvaro Obeso Fernandez, MSc, Helsinki Institute for Demography and Public Health, University of Helsinki Unioninkatu 33, 00170 University of Helsinki Finland, GSM.

## Abstract

**Introduction:** Educational attainment (EA) is negatively associated with body mass index (BMI) but less is known about the association between EA and adult BMI change. We analyzed the role of genetic and environmental factors in the association between EA and BMI trajectory components over adulthood.

**Data and methods:** Pooled data from 59,490 twins aged 31–99 years (49% women) across 11 cohorts with EA and repeated measures of BMI were used. BMI trajectory components (baseline BMI and BMI change per decade) were estimated using linear mixed-effects (LME) and delta slope methods. EA was derived by regressing years of education on birth year and cohort. Associations between EA and BMI trajectories were evaluated with LME models in both cohort-specific and pooled data. Genetic and environmental contributions were evaluated using structural equation modeling.

**Results:** EA was more strongly negatively associated with baseline BMI and BMI change (mean of 1.31 and 1.32 kg/m^2^ per decade in men and women, respectively) in women (β = – 0.14 kg/m², 95% CI: –0.15 to –0.12; β = –0.02 kg/m²/decade, 95% CI: –0.03 to –0.01, respectively) than in men (β = –0.07, 95% CI: –0.08 to –0.06; β = –0.01, 95% CI: –0.02 to – 0.001, respectively). The associations between baseline BMI and EA were explained by genetic factors in men (r_A_ = –0.10) and by both genetic (r_A_ = –0.17) and unique environmental factors (r_E_ = –0.07) in women. For BMI change, the associations with EA were explained solely by genetic factors (r_A_ = –0.04 in men; –0.06 in women).

**Conclusion:** Individuals with higher EA tend to have lower baseline BMI and slower BMI increases across adulthood. The majority of the associations are primarily genetically mediated.

## Introduction

Educational attainment (EA) is a key social indicator in epidemiology (1), influencing health behaviors, occupational status, and lifetime earnings (2). A large body of research has documented strong associations between EA and various health outcomes (3, 4) as well as mortality (5). One important health outcome linked to EA is obesity, which is typically measured using body mass index (BMI) in epidemiological studies as it strongly reflects fat mass and body fat percentage at the population level (6). Studies across diverse populations have shown that higher personal EA is associated with lower adult BMI (7–10) and that parental EA also influences offspring’s BMI in childhood and adulthood (11, 12). Obesity prevalence has nearly tripled over the past 40 years (13, 14), potentially reinforcing educational health inequalities since BMI impacts on multiple health outcomes (15–17) and mortality (16).

Both genetic and environmental factors influence EA and obesity. The development of obesity is affected by developmental, environmental, behavioral, and genetic factors, as well as their mutual interactions (18, 19, 20). Previous twin studies have reported that from half to nearly 90% of individual differences in adult BMI are attributed to genetic differences (21, 22, 23). This strong genetic influence has also been supported by several genome-wide association studies (GWAS), which have identified numerous genetic variants associated with BMI variation in both childhood (24) and adulthood (25, 26) through cross-sectional analyses. For BMI change, genetic factors appear to play a less important role. Previous studies have reported only slightly moderate correlations (0.25–0.27) between BMI change and polygenic risk scores of BMI (27). More recently, another study found the heritability of BMI change to be low across different stages of adulthood, remaining below 30% in both men and women at all stages (28). For EA, large-scale twin studies have reported that from a third to half of individual difference in EA are due to genetic differences (29). GWAS have identified multiple single-nucleotide polymorphisms (SNPs) associated with EA, collectively explaining between 3% and 16% of its variance (30, 31). Furthermore, there is evidence of genetic correlations between EA and BMI based on twin (-0.20 in men and women) (32) and population-based studies (-0.20 to -0.25) (33).

Although the association between EA and BMI has been studied cross-sectionally, few studies have examined EA in relation to adult BMI change. Furthermore, the genetic and environmental contributions to these associations remain unclear. We aimed to address these gaps by analyzing the phenotypic associations between EA and BMI trajectory components (Baseline BMI and BMI change) across adulthood, as well as the underlying genetic and environmental influences on these associations, using longitudinal data from 11 twin cohorts across 8 countries.

## Data and methods

### Cohort

This study used data from the CODATwins project, which combined twin cohorts worldwide with height and weight information, in collaboration with the BETTER4U consortium (34, 35). Additional data on EA were available for some cohorts (36). Thirteen cohorts with EA and longitudinal BMI data collected after age 18 were included: seven from Europe, four from the USA, and one each from Australia and South Korea, with two cohorts consisting only of men. Cohorts were categorized into four regions: Europe, North America, East Asia, and Australia. BMI was calculated as weight divided by height squared (kg/m²) using self-reported (89%) or measured (11%) values. The original education categories were converted to years of education by assigning the mean years for each category and then harmonized by regressing education years on birth year and cohort (36). The residuals from this model represent individual deviations from the expected education level given birth year and cohort.

Initially, the database included 88,773 twin individuals with data on EA and at least two measures of BMI. From this data, we excluded individuals who reported their EA before 30 years old to ensure they had completed their education (N= 24,922). Subsequently, the distributions of baseline BMI (Supplementary Figure 1) and BMI change (Supplementary Figure 2) were examined using scatter plots. Individuals with values greater than 3 standard deviations from the mean were considered outliers and removed prior to analysis (N = 2,400 for baseline BMI and N = 1,961 for BMI change). As a result, a final sample of 59,490 twins (49% women) and 22,129 complete twin pairs (44% monozygotic (MZ), 47% same-sex dizygotic (DZ) and 9% opposite-sex DZ) belonging to 11 cohorts was obtained. The participant selection process is summarized in Supplementary Figure 3.

### BMI trajectory components calculation

BMI trajectory components were derived using two methods based on the number of BMI measurements. For participants with only two measurements, the delta method was applied, calculating the difference between BMI values divided by the time elapsed. For those with three or more measurements, linear mixed-effects (LME) models were used, which account for both fixed and random effects and the correlation between repeated observations (37). Both approaches provided baseline BMI and the rate of BMI change for each participant, with LME models estimating individual-level variation through random effects on these trajectory components. BMI change was calculated as annualized change and scaled to per decade to allow comparison across participants with different follow-up times. Adult BMI change was assumed to be approximately linear between ages 18 and 60, so the annualized slope reflects each individual’s typical rate of BMI change.

### Association analysis

Associations between BMI trajectory components and EA were examined using LME models at both cohort-specific and pooled levels. All analyses were stratified by sex, given the known sex differences in BMI trajectories (23, 27, 38).

In cohort-specific analyses, baseline BMI was modeled as the dependent variable and EA as the independent variable, adjusting for baseline age and zygosity. BMI change was then modeled similarly, additionally including baseline BMI as a covariate. Heterogeneity across cohorts was assessed using Cochran’s Q test, which evaluates whether observed differences in effect estimates are compatible with chance under a common effect assumption (39).

When using the pooled data, a cohort-specific identifier was included as a covariate in each of the models described above to account for between-cohort differences. We tested the linearity of EA associations with baseline BMI and BMI change by including quadratic age terms and applying flexible models using restricted cubic splines and generalized additive models (GAMs). In all LME models, random intercepts for family ID were included to account for the non-independence of observations due to familial relatedness.

Two sensitivity analyses were conducted. First, smoking behavior at baseline (classified as current smokers, never smokers, and former smokers) was included as a covariate (N = 15,002), given its well-established association with EA (40, 41, 42) and with BMI and weight change (43, 44, 45). Smoking was treated as a potential confounder, as it could influence both EA and BMI trajectory components. Participants with missing data on smoking, BMI trajectory components, or EA were excluded without imputation. Secondly, a sensitivity analysis dividing adulthood into stages based on the first and last BMI measurement (middle to late adulthood: 30–50 years; middle age: 51–65 years; old age: >66 years) was carried out to examine age-specific associations, as BMI trajectories vary across adulthood (28, 46). All statistical analyses were performed using R software (version 4.2.3) with the lme4 package (version 1.1-34). The Cochran’s Q tests were conducted using the metafor (version 3.8.1) and meta (version 6.3.0) R packages.

### Structural equation modeling

Data were analyzed using structural equation modeling (SEM), which leverages differences in genetic relatedness between MZ twins who are genetically virtually identical at the gene sequence level and DZ twin sharing 50% of genes identical-by-descent (47). Trait variance was decomposed into additive genetic variance (A; correlation 1 in MZ, 0.5 in DZ), dominance genetic variance (D; correlation 1 in MZ, 0.25 in DZ), shared environmental variance (C; correlation 1 in both MZ and DZ), and unique environmental variance including also measurement error variance (E; correlation 0 in both MZ and DZ). Since D and C cannot be estimated simultaneously with only twins reared together, either an additive genetic/ shared environmental/unique environmental (ACE) or additive genetic/ dominance genetic/unique environmental (ADE) model was specified and tested for simplification to an AE model. Consistency of intraclass correlations (ICC) across cohorts was used to justify pooling. The baseline model (ACE or ADE) was first selected using ICCs calculated as the covariance between twins divided by the total variance of the trait, and their 95% confidence intervals (CI) were obtained based on the F distribution (48). The baseline model was then compared to the more parsimonious AE model. The best-fitting model was subsequently tested against submodels excluding (i) sex differences and (ii) sex-specific genetic effects using the χ²-distributed –2LL test.

A bivariate Cholesky decomposition was conducted to investigate the genetic and environmental contributions to the significant associations between EA and BMI trajectory components, both at the cohort-specific level—to assess potential cohort differences—and in the pooled data to maximize power. Cholesky decomposition is a model-free approach that decomposes the variance and covariance between two traits into additive genetic (A), shared environmental (C), and unique environmental (E) components via independent latent factors (49). Standardized genetic and environmental correlations were calculated by dividing the covariance of each component by the square root of the product of the corresponding trait variances, yielding genetic (r_A_), shared environmental (r_C_), and unique environmental (r_E_) correlations (49). Confidence intervals (95% CI) were estimated by maximum likelihood (49). At the cohort-specific level, heterogeneity of the results was assessed by Cochran’s Q test (39). Sex differences were modeled by creating separate SEM subsets for male-male (MZM, DZM) and female-female (MZF, DZF) twin pairs. Opposite-sex DZ (OSDZ) pairs were included with a freely estimated genetic correlation to contribute consistently to all SEM analyses. Analyses were performed in R (version 4.2.3) using OpenMx (version 2.21.11), and Cochran’s Q tests were conducted with the metafor (version 3.8.1) and meta (version 6.3.0) packages.

## Results

Descriptive characteristics of the cohorts included in the study after applying exclusion criteria are shown in Table 1. The Vietnam Era Twin Study of Aging had the highest baseline BMI (29.07 kg/m²) and the oldest baseline age (56.29 years), whereas the Netherlands Twin Registry had the lowest baseline BMI (21.62 kg/m²) and youngest baseline age (31.26 years). The greatest BMI change per decade was observed in the Washington State Twin Registry (1.40 kg/m²), while the lowest was seen in the Colorado Twin Registry and Korea Twin-Family Register (0.60 kg/m²). EA was highest in the Washington State Twin Registry (15.26 years) and lowest in the Finnish Old Cohort (8.05 years).The mean follow up time was 15.83 years, being NAS-NCR Twin registre the one with highest follow up (22.60 years) and Italian Twin Registry the one with lowest follow up time (4.23 years).

**Table 1.** Descriptive information of the cohorts included in the database after the application of exclusion criteria. Descriptive information of the 13 cohorts that form the CODATwins database used in the current study after application of exclusion criteria**. Abbreviations**: N: number; SD: Standard deviation; NAS-NRC: National Academy of Sciences-National Research Council.

| Geographic area and cohorts | General information |  |  | Follow up information |  | Baseline information |  |  | Longitudinal information |  | Information on education |  |
| --- | --- | --- | --- | --- | --- | --- | --- | --- | --- | --- | --- | --- |
|  | Age range | N of individuals | % women | N of measures | Mean follow-up time (years) | Mean baseline age (years) | Mean Baseline BMI | Mean birth year | BMI change per ten years |  | Mean education years |  |
|  |  |  |  |  |  |  |  |  | Mean | SD | Mean | SD |
| *Europe* |  |  |  |  |  |  |  |  |  |  |  |  |
| Finnish old cohort | 31–100 | 20,939 | 53% | 4 | 19.21 | 33.65 | 23.10 | 1951 | 0.80 | 0.40 | 8.05 | 3.29 |
| Italian Twin Registry | 31–78 | 1,280 | 67% | 4 | 4.23 | 34.19 | 22.27 | 1973 | 0.80 | 3.40 | 13.40 | 2.26 |
| Murcia Twin Register | 31–89 | 1,298 | 64% | 8 | 7.39 | 35.97 | 25.84 | 1963 | 0.80 | 0.70 | 10.67 | 3.91 |
| Netherlands Twin Register | 31–91 | 3,961 | 66% | 13 | 10.65 | 31.26 | 21.62 | 1968 | 0.90 | 0.40 | 13.96 | 2.45 |
| Swedish Twin Register | 31–99 | 8,255 | 46% | 4 | 19.90 | 39.77 | 24.51 | 1940 | 1.00 | 0.90 | 11.96 | 3.08 |
| *North America* |  |  |  |  |  |  |  |  |  |  |  |  |
| Colorado Twin Registry | 31–34 | 1,478 | 56% | 3 | 6.62 | 31.69 | 22.68 | 1985 | 0.60 | 0.40 | 14.51 | 2.17 |
| NAS-NRC Twin Registry | 31–82 | 8,692 | 0% | 4 | 22.60 | 44.26 | 22.22 | 1923 | 0.80 | 0.60 | 13.52 | 3.09 |
| Vietnam Era Twin Study of Aging | 51–66 | 743 | 0% | 2 | 5.76 | 56.29 | 29.07 | 1950 | 0.90 | 3.70 | 13.88 | 2.09 |
| Washington State Twin Registry | 31–97 | 5,877 | 66% | 6 | 6.67 | 35.67 | 24.81 | 1970 | 1.40 | 5.70 | 15.26 | 2.38 |
| *Asia* |  |  |  |  |  |  |  |  |  |  |  |  |
| Korea Twin-Family Register | 31–79 | 200 | 65% | 4 | 4.37 | 38.73 | 23.07 | 1968 | 0.60 | 3.00 | 14.24 | 3.92 |
| *Australia* |  |  |  |  |  |  |  |  |  |  |  |  |
| Queensland Twin Register | 31–92 | 6,767 | 51% | 9 | 14.00 | 33.18 | 22.75 | 1959 | 1.10 | 4.20 | 15.16 | 2.07 |
| *World* |  |  |  |  |  |  |  |  |  |  |  |  |
| All cohorts | 31–100 | 59,490 | 49% | 2-13 | 15.83 | 37.89 | 22.54 | 1952 | 0.90 | 2.60 | 11.81 | 4.15 |

Figure 1 shows the cohort-specific associations between EA and baseline BMI, while Figure 2 presents the results for BMI change over a decade (the exact estimates are available in Supplementary Table 1). Higher EA was generally linked to lower baseline BMI and smaller BMI increases. Statistically significant associations for baseline BMI were observed in five cohorts of men and eight cohorts of women, and for BMI change in two cohorts of men and four cohorts of women. Cochran’s Q tests indicated significant heterogeneity across cohorts for all outcomes in both sexes (all p < 0.001).

**Figure 1.**
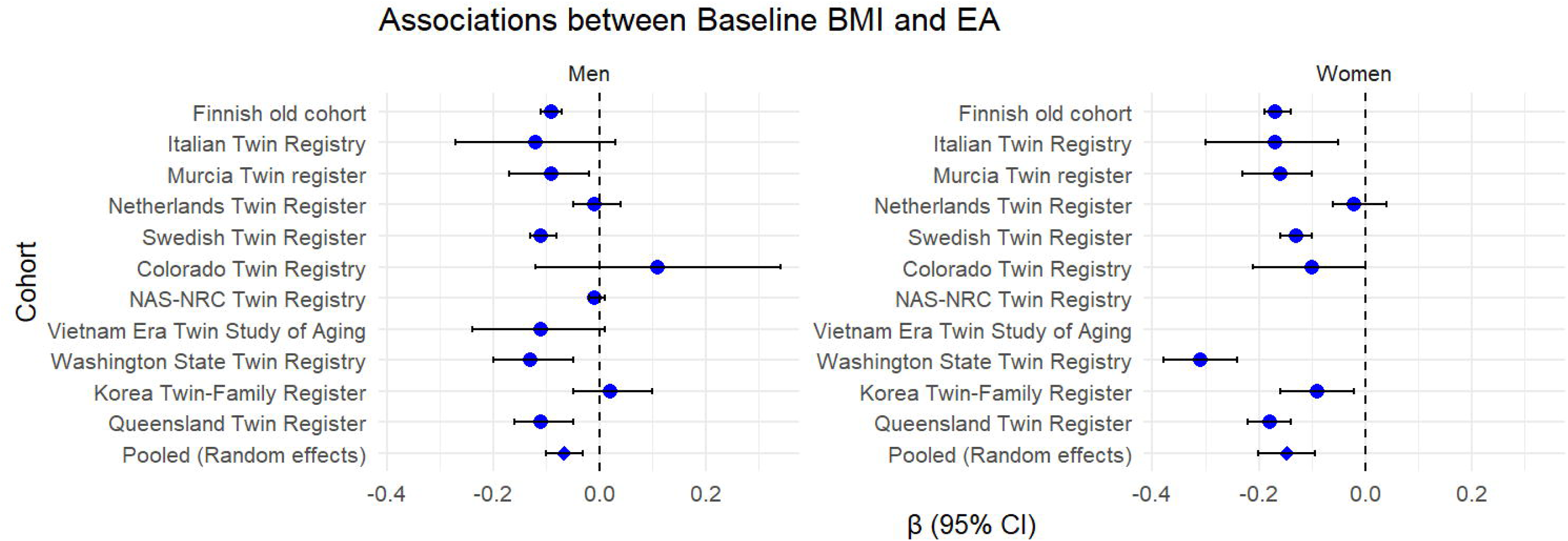
Association between educational attainment and baseline BMI by sex (left in men and right in women) and twin cohort. Forest plots showing the association between educational attainment and baseline BMI in men and women across multiple twin cohorts. Each dot represents the point estimate of the association for a specific cohort, with horizontal lines indicating 95% confidence intervals. Cochran’s Q test indicates significant heterogeneity across cohorts (Men: Q = 120.97, p = 3.95e-20; Women: Q = 125.38, p = 4.06e-22). **Abbreviations:** BMI: body mass index.

**Figure 2.**
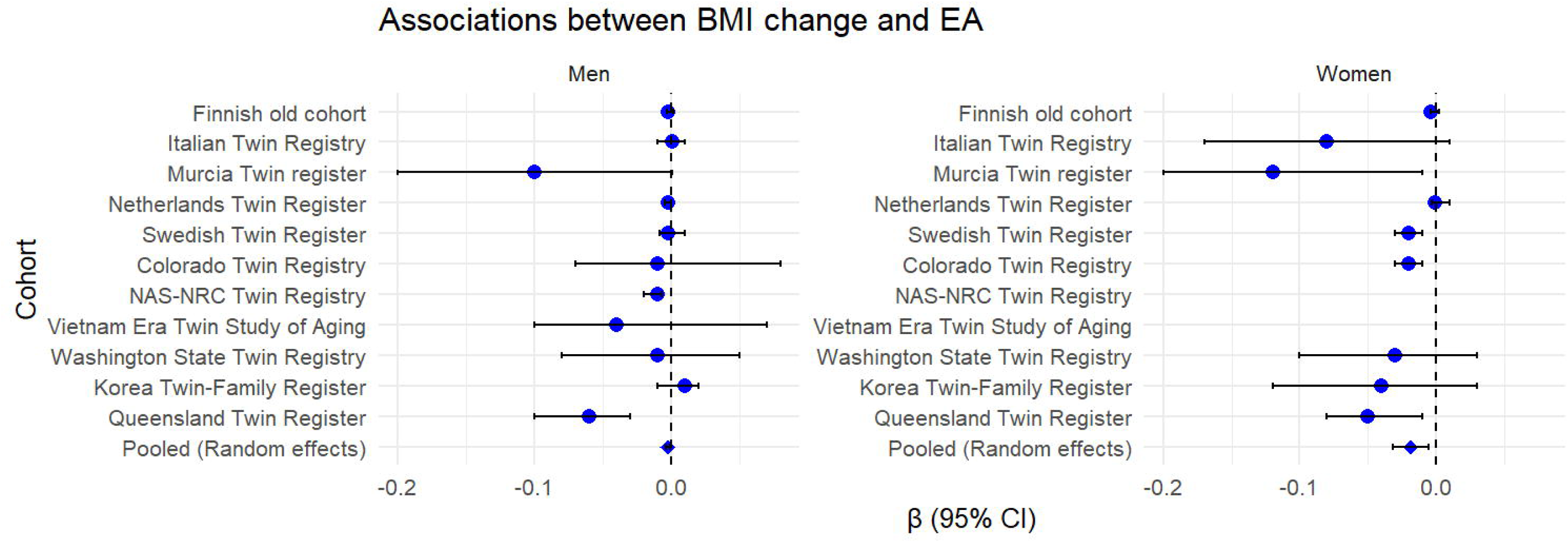
Association between educational attainment and BMI change by sex (left in men and right in women) and twin cohort. Forest plots showing the association between educational attainment and BMI change in men and women across multiple twin cohorts. Each dot represents the point estimate of the association for a specific cohort, with horizontal lines indicating 95% confidence intervals. Cochran’s Q test indicates significant heterogeneity across cohorts (Men: Q = 42.88, p = 2.35e-05; Women: Q = 65.00, p = 4.04e-10). **Abbreviations:** BMI: body mass index.

The results of the associations of EA with baseline BMI and BMI change using pooled data (N= 59,490) are displayed in Table 2. EA was negatively associated with baseline BMI and BMI change in both men and women. In men, a one standard deviation (SD) higher EA (approximately 3 years) was associated with a 0.07 kg/m² lower baseline BMI and a 0.01 kg/m² per decade slower increase in BMI. In women, a one SD higher EA (2.80 years) was associated with a 0.14 kg/m² lower baseline BMI and a 0.02 kg/m² per decade slower rate of BMI change. Linearity tests confirmed the robustness of associations.

**Table 2.** Association study of EA with baseline BMI (kg/m^2^) and BMI change (kg/m^2^ per ten year) in pooled data, by sex. Associations analyses were carried out using LME models. The models are: i) baseline BMI∼ EA + cohort ID + Zygosity + Baseline Age + (1|Family ID) and ii) BMI change∼ EA + cohort ID + Zygosity + Baseline Age + baseline BMI + (1|Family ID). Variables were harmonized prior to analysis, and results are interpreted as effect sizes per 1 SD change in EA. The estimates are displayed along with the 95% confidence interval. **Abbreviations:** BMI: Body mass index; CI: confidence interval; LL: lower limit; UL: upper limit

| Variables of study | Men (N=33,763) |  |  | Women (N=31,314) |  |  |
| --- | --- | --- | --- | --- | --- | --- |
| | $\beta$ | 95%CI | | $\beta$ | 95%CI | |
|  |  | LL | UL |  | LL | UL |
| Baseline BMI | -0.07 | -0.08 | -0.06 | -0.14 | -0.15 | -0.12 |
| BMI change | -0.01 | -0.02 | -1.26e-03 | -0.02 | -0.03 | -0.01 |

LME models detected a significant quadratic age effect on BMI slope (p < 0.001) but no significant EA–age interactions (men: p > 0.05; women: p = 0.167 and 0.648). GAMs showed non-linear age effects (men: edf = 3.47, F = 38.91, p < 0.001; women: edf = 4.72, F = 27.15, p < 0.001) without significant EA–age interactions (men: p = 0.343; women: p = 0.277). Restricted cubic spline models yielded consistent EA effects, with only minor late-age interactions that did not improve model fit (men: χ² = 6.34, df = 4, p = 0.175; women: χ² = 7.53, df = 4, p = 0.111). The sensitivity analyses results—including (i) adjustment for smoking and (ii) stratification of adulthood into stages—are shown in Supplementary Tables 2 and 3. Associations remained largely significant after adjusting for smoking (N=15,002) (Supplementary Table 2). When stratified by adulthood stage (N= 19451, 2,892 and 545 individuals in middle to late adulthood, middle age and old age, respectively), the significance of associations varied across men and women depending on specific stage (Supplementary Table 3).

**Table 3.** Relative proportions of the variability of baseline BMI, BMI change and EA explained by additive genetic, shared environmental and unique environmental variance components with 95% confidence intervals. Full AE model was selected for baseline BMI and BMI change, while full ACE model for EA. **Abbreviations:** a^2^: proportion of total variance explained by additive genetic effects; c^2^: proportion of total variance explained by shared environmental effects; e^2^: proportion of total variance explained by unique environmental effects; LL: Lower limit; UL: Upper limit.

| Variables of study | Additive genetic effect |  |  | Shared environmental effect |  |  | Unique environmental effect |  |  |
| --- | --- | --- | --- | --- | --- | --- | --- | --- | --- |
| | $a^2$ | 95% CI | | $c^2$ | 95% CI | | $e^2$ | 95% CI | |
|  |  | LL | UL |  | LL | UL |  | LL | UL |
| Men |  |  |  |  |  |  |  |  |  |
| Baseline BMI | 0.81 | 0.79 | 0.82 | - | - | - | 0.19 | 0.18 | 0.20 |
| BMI change | 0.21 | 0.17 | 0.23 | - | - | - | 0.79 | 0.76 | 0.82 |
| EA | 0.41 | 0.37 | 0.45 | 0.30 | 0.26 | 0.33 | 0.29 | 0.27 | 0.30 |
| Women |  |  |  |  |  |  |  |  |  |
| Baseline BMI | 0.79 | 0.78 | 0.80 | - | - | - | 0.21 | 0.20 | 0.22 |
| BMI change | 0.19 | 0.16 | 0.21 | - | - | - | 0.81 | 0.78 | 0.83 |
| EA | 0.31 | 0.27 | 0.35 | 0.42 | 0.38 | 0.44 | 0.27 | 0.26 | 0.28 |

We began genetic modeling by selecting the best-fitting model based on MZ and DZ ICCs (Supplementary Table 4). The ACE model was selected as the baseline model because MZ correlations were less than twice DZ correlations. Removing the shared environment variance did not impact the model fit for baseline BMI or BMI change, but it did affect EA (Supplementary Table 5). No sex-specific genetic effects were detected. Therefore, we used the AE model for BMI and an ACE model for EA in the subsequent analyses, both without qualitative sex differences but including sex-specific variance components.

The results of the genetic and environmental factors influencing the variation of baseline BMI, BMI change, and EA are displayed in Table 3. For baseline BMI, the genetic additive effect explained most of the variation in men (a^2^ = 0.81) and women (a^2^ = 0.79). The variation in BMI change was mainly explained by unique environmental factors in men (e^2^ = 0.79) and women (e^2^ = 0.81). For EA, additive genetic effects explained a substantial part of the variation in men (a^2^ = 0.41) followed by shared environmental effects (c^2^ = 0.30), while in women, shared environmental effects explained most of the variation (c^2^ = 0.42) followed by additive genetic effects (a^2^ = 0.31).

The associations between BMI trajectory components and EA were explained by genetic and unique environmental correlations (Table 4). The shared environment effect was not considered since it was found only for EA and thus cannot explain these correlations. In men, the association between EA and baseline BMI was primarily driven by additive genetic factors (r_A_ = –0.10), whereas in women, it was explained by both additive genetic (r_A_ = – 0.17) and unique environmental factors (r_E_ = –0.07). For BMI change per decade, EA associations were entirely attributable to additive genetic factors in both men (r_A_ = –0.04) and women (r_A_ = –0.06). The cohort-specific results from bivariate Cholesky decomposition are shown in Figures 3 and 4 (the exact estimates are available in Supplementary Table 6). The contributions of genetic and environmental factors varied across the cohorts. Cochran’s Q tests indicated significant heterogeneity for r_A_ and r_E_ in baseline BMI in both sexes (all p<0.001). For BMI change, heterogeneity was found only for r_E_ in men (p<0.001) and r_A_ in women (p = p<0.001).

**Figure 3.**
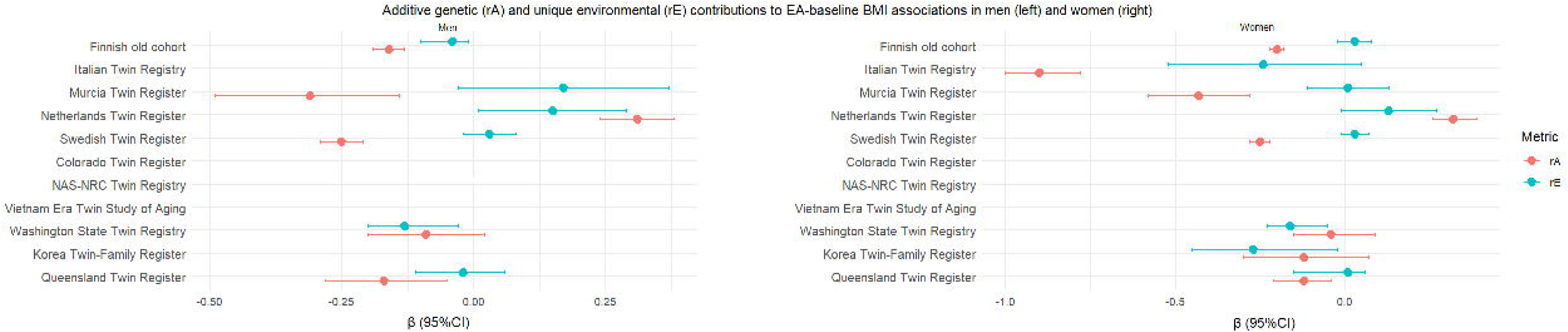
Cohort specific additive genetic (r_A_) and unique environmental (r_E_) correlations underpinning the associations between baseline BMI and EA in men (left) and women (right). Forest plot showing estimates of genetic (r_A_, red) and unique environmental (r_E_, teal) correlations obtained from the associations between BMI baseline BMI and EA across multiple cohorts in men in the left and women in the right. Points represent correlation estimates, and horizontal lines represent 95% confidence intervals. Cochran’s Q tests indicate significant heterogeneity for baseline BMI (r_A_: Q = 195.14, p = 2.05e-39; r_E_: Q = 21.27, p = 3.38e-03) and BMI change (r_A_: Q = 1.76, p = 0.62; r_E_: Q = 14.72, p = 2.06e-03). **Abbreviations:** BMI: body mass index; CI: confidence interval.

**Figure 4.**
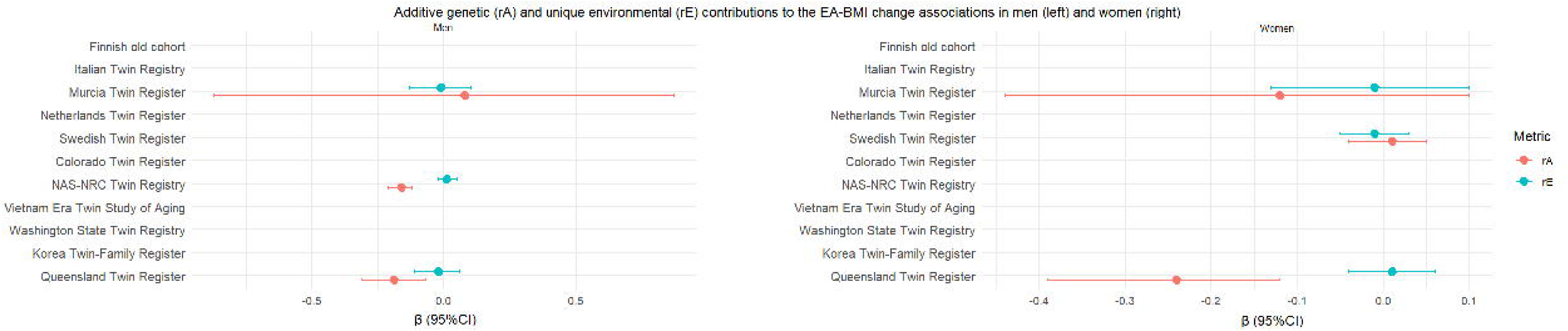
Cohort specific additive genetic (r_A_) and unique environmental (r_E_) correlations between BMI change and EA in men (left) and women (right). Forest plot depicting cohort-level estimates of additive genetic (r_A_, red) and unique environmental (r_E_, teal) correlations between BMI change and EA in men in the left and women in the right. Each point represents a correlation estimate, with horizontal lines indicating 95% confidence intervals. Cochran’s Q tests indicate significant heterogeneity for baseline BMI (r_A_: Q = 441.93, p = 1.52e-89; r_E_: Q = 46.10, p = 1.37e-06) and BMI change (r_A_: Q = 0.49, p = 0.92; r_E_: Q = 12.59, p = 5.58e-03). **Abbreviations**: BMI: body mass index; CI: confidence interval.

**Table 4.** Contribution of additive genetic and unique environmental factors to the previously identified associations between BMI trajectory components (baseline BMI and BMI change) and EA by sex. Additive genetic (r_A_) and unique environmental (r_E_) correlations of EA with baseline BMI and BMI change were obtained from structural equation models, which decompose phenotypic covariation into genetic and environmental effects. **Abbreviations**: r_A_: Additive genetic correlation coefficient; r_E_; Specific environmental correlation coefficient; LL: Lower limit; UL: Upper limit.

| Variables of study | Men |  |  |  |  |  | Women |  |  |  |  |  |
| --- | --- | --- | --- | --- | --- | --- | --- | --- | --- | --- | --- | --- |
|  | Additive genetic correlation |  |  | Unique environmental correlations |  |  | Additive genetic correlation |  |  | Unique environmental correlation |  |  |
| | $r_A$ | 95% CI | | $r_E$ | 95% CI | | $r_A$ | 95% CI | | $r_E$ | 95% CI | |
|  |  | LL | UL |  | LL | UL |  |  |  |  |  |  |
| Baseline BMI | -0.10 | -0.12 | -0.08 | -0.01 | -0.04 | 0.01 | -0.17 | -0.19 | -0.15 | -0.07 | -0.09 | -0.05 |
| BMI change | -0.04 | -0.07 | -0.01 | -0.01 | -0.03 | 0.01 | -0.06 | -0.10 | -0.02 | -0.01 | -0.03 | 0.01 |

## Discussion

Our findings showed that EA was negatively associated with baseline BMI and BMI change both in men and women. Furthermore, genetic and environmental factors explained the variation in these variables as well as the associations between them. Individuals with higher education level tend to have lower baseline BMI and slower BMI increases across adulthood. However, when adulthood was divided into three stages, the associations with baseline BMI and BMI change varied by age and sex: men showed significant associations only for baseline BMI in middle to late adulthood (30–50 years), whereas women had significant associations with baseline BMI across all stages (with the strongest association in old age), and with BMI change up to old age (with the strongest association in middle age). This pattern may reflect reduced statistical power in the stratified analyses but may also indicate that the protective effect of education on BMI trajectories differs across adulthood — particularly among women—and that this effect is especially pronounced during the middle and later stages of adulthood.

These results align with previous studies showing a higher risk of obesity among individuals with lower EA (50–52). Longitudinal research has further found that those with higher EA tend to gain less weight over time and are less likely to transition into overweight or obese categories during adulthood (53, 54). EA may also moderate the impact of socioeconomic disadvantage on obesity, potentially buffering against weight gain in low-income groups by promoting protective health behaviors (55). Differences in health literacy, dietary patterns, and physical activity associated with EA have been suggested as key mechanisms underlying this relationship (56, 57). However, to our knowledge, no prior research has specifically examined the contributions of genetic and environmental factors to the association between EA and BMI change over time.

Cohort-specific results revealed substantial heterogeneity in the strength and significance of the associations between BMI trajectories and EA, reflecting differences in sample size, age distribution, and follow-up duration. Larger cohorts showed consistent negative associations between baseline BMI and EA, whereas smaller cohorts displayed wider confidence intervals and less consistent patterns, particularly for BMI change. Temporal and cultural factors likely contribute to these differences, as variations in public health policies, nutritional environments, and socioeconomic conditions can modulate how education translates into health behaviors and BMI outcomes, even though identifying features of an obesogenic environment is challenging (58). For example, earlier-born cohorts may have experienced different educational opportunities and dietary environments than younger cohorts. Similarly, European cohorts with longer follow-up may capture more gradual BMI trajectories, whereas smaller or shorter-follow-up cohorts may have less power to detect change. Differences in data collection timing further complicate comparisons, making it difficult to disentangle age, period, and country-level effects.

Genetic twin modeling revealed distinct sources of individual variance for the traits examined. For baseline BMI, additive genetic effects were the primary contributors to individual differences in both men and women, with heritability estimates (**∼** 0.80) closely matching those previously reported in early adulthood using cross-sectional BMI data from the CODATwin project (20). In contrast, variability in BMI change was predominantly explained by unique environmental factors, highlighting the greater influence of individual-specific experiences and exposures, as observed in prior CODATwin analyses with larger cohort samples (28). The higher proportion of variance attributable to unique environmental factors in women may partly reflect weight gain following pregnancy contributing to greater environmental variance. For EA, sources of variance differed by sex: in men, additive genetic effects were the main contributor, followed by shared environmental effects, which slightly exceeded unique environmental influences. Conversely, in women, shared environmental effects contributed most strongly to EA variance, followed by additive genetic effects, consistent with previous findings (29).

Our key findings pertain to the underlying factors linking EA and BMI change. In men, the associations of EA with both baseline BMI and BMI change were fully explained by negative associations attributable to additive genetic factors, suggesting that genetic predispositions linked to higher education also contribute to lower BMI and slower BMI increase. In women, the association between baseline BMI and EA was driven by both additive genetic and unique environmental factors, while the association between BMI change and EA was explained exclusively by additive genetic factors. These findings suggest that, although education is often linked to healthier lifestyles and weight stability, the associations between EA and both baseline BMI and BMI change are primarily genetically mediated. The small unique environmental contribution observed in women for baseline BMI indicates that the observed relationships are unlikely to reflect a causal effect of education on BMI, at least in men, consistent with the non-significant environmental correlations in our bivariate twin analyses. Ultimately, this points to potential sex differences in the interplay between educational experiences and biological pathways shaping long-term health outcomes. Moreover, cohort-specific analyses indicated that the relative contribution of genetic and environmental factors varied across populations, reflecting differences in social, cultural, and healthcare context and potentially also differences in age groups and/or follow-up periods. While genetic influences were generally consistent, the role of unique environmental factors appeared more variable.

Overall, these findings underscore that the interplay between genetic and environmental factors in shaping BMI trajectories is context-dependent, and that the protective influence of education on BMI may operate differently depending on both environmental exposures and cohort-specific characteristics.

Our findings underscore the importance of considering both sex and cohort-specific contexts when interpreting the relationship between socioeconomic factors and long-term health trajectories. A previous study examining genetic and environmental contributions to education-related differences in BMI and weight change found that the negative correlation between baseline BMI and EA was largely driven by additive genetic factors, with only a modest positive unshared environmental effect observed in women. Additionally, that study reported that the same genetic factors influencing baseline BMI and EA also contributed to the EA–weight change association (32). To our knowledge, however, no prior research has examined the genetic and environmental underpinnings of associations between EA and BMI trajectory components. Our results extend the literature by applying a longitudinal, trajectory-based approach, enabling the decomposition of genetic and environmental contributions not only to baseline BMI but also to patterns of BMI change over time.

The current study has several strengths, most notably the inclusion of a large sample of twins from multiple cohorts with EA and longitudinal BMI data. Additionally, information on smoking status allowed us to account for this important confounder, enhancing the validity of the observed associations between BMI trajectory components and EA. This comprehensive dataset strengthens our ability to disentangle genetic and environmental contributions over time and across individuals. However, there are also limitations. First, the absence of data from low- and middle-income countries limits the generalizability of our findings. Second, our analyses include only completed levels of education; incorporating additional factors such as academic performance, cognitive ability, motivation, and other influences on educational outcomes would provide deeper insight into how BMI and its changes relate to EA and the genetic and environmental mechanisms underlying these associations. Assortative mating—where partners are chosen based on similarity in observable or known traits—can affect EA, a highly heritable trait (29), by increasing the genetic correlation between spouses. This can raise the additive genetic correlation of DZ twins and full siblings above the expected 0.5 under random mating, potentially inflating shared environmental estimates and biasing the separation of genetic and environmental contributions in classical twin designs (59, 60). In this study, accounting for assortative mating was not possible due to missing spouse information in several cohorts. Finally, while smoking data were available, information on other potential modifiers—such as nutrition, physical activity, childbirth, and lifestyle factors—was lacking. These factors could also influence EA and BMI trajectories and may contribute to the observed sex differences.

In conclusion, this study shows that the associations between EA and BMI trajectories across adulthood are primarily driven by additive genetic factors, with minimal influence from individual-specific environmental factors. EA is negatively related to both baseline BMI and BMI change, with genetic factors exclusively underlying these associations in men, and both genetic and unique environmental factors contributing in women, particularly for baseline BMI. These findings highlight the value of dynamic weight measures and reveal important sex-specific differences in the etiology of BMI and its relationship with education. While the results are based on a large twin sample from multiple cohorts, which enhances robustness, generalizability may be somewhat limited to populations with similar socio-cultural and educational contexts

## Ethics statement

The pooled analysis was approved by the ethical board of the Department of Public Health, University of Helsinki. The data collections procedures of participating twin cohorts were approved by local ethical boards following the regulation in each country. Only anonymized data were delivered to the data management center at University of Helsinki.

## Data availability

The data used in this study is owned by the third parties (the individual twin cohorts) and made available to this study in condition that they will be used only in this meta-analysis. The data can be used based on the same principles as used in this study (more information from Karri Silventoinen).

## Code availability

All R scripts are available from the corresponding author.

## Acknowledgment

AO and KS designed the study. EM, CF, VT, AL, SA, SM, SG, JL, SL, JS, HP, GD, DB, JO, JSR, LCC, CF, WK, RC, BH, PM, IK, ADA, ML, MB, LL, EG, MG, DB, NM, DIB, JK collected the original data files. KS, JK, AJ and AO pooled the data together. GD developed part of the scripts for the analysis. AO conducted the analyses and drafted the manuscript. All authors revised the manuscript, approved the final version and agreed to be accountable for all aspects of the work in ensuring that questions related to the accuracy or integrity of any part of the work are appropriately investigated and resolved.

