## Supplementary material for "Genetic and environmental influences on educational disparities in adult weight change: an individual-based pooled analysis of 11 twin cohorts"

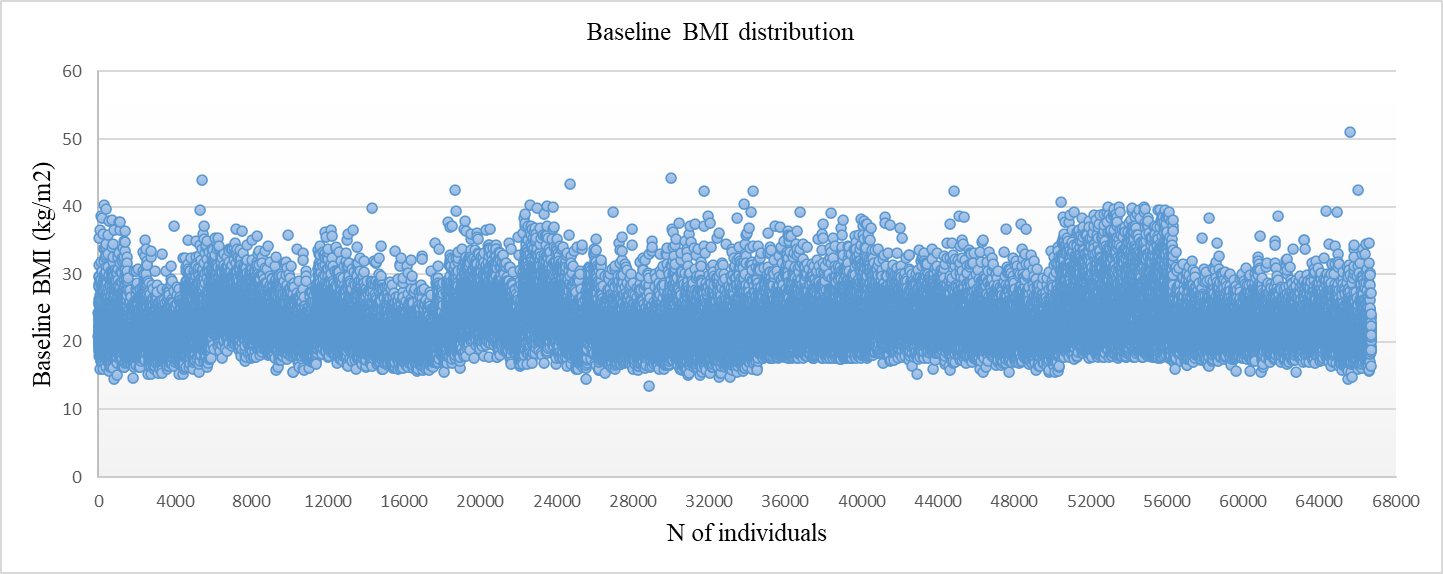
**Supplementary Figure 1**. Baseline BMI distribution for identifying outliers.

**Caption:** Scatter plot displaying baseline BMI values is shown in the figure. Individuals with BMI values greater than 3 standard deviations from the mean were considered outliers and removed prior to analysis. **Abbreviations:** BMI: body mass index.

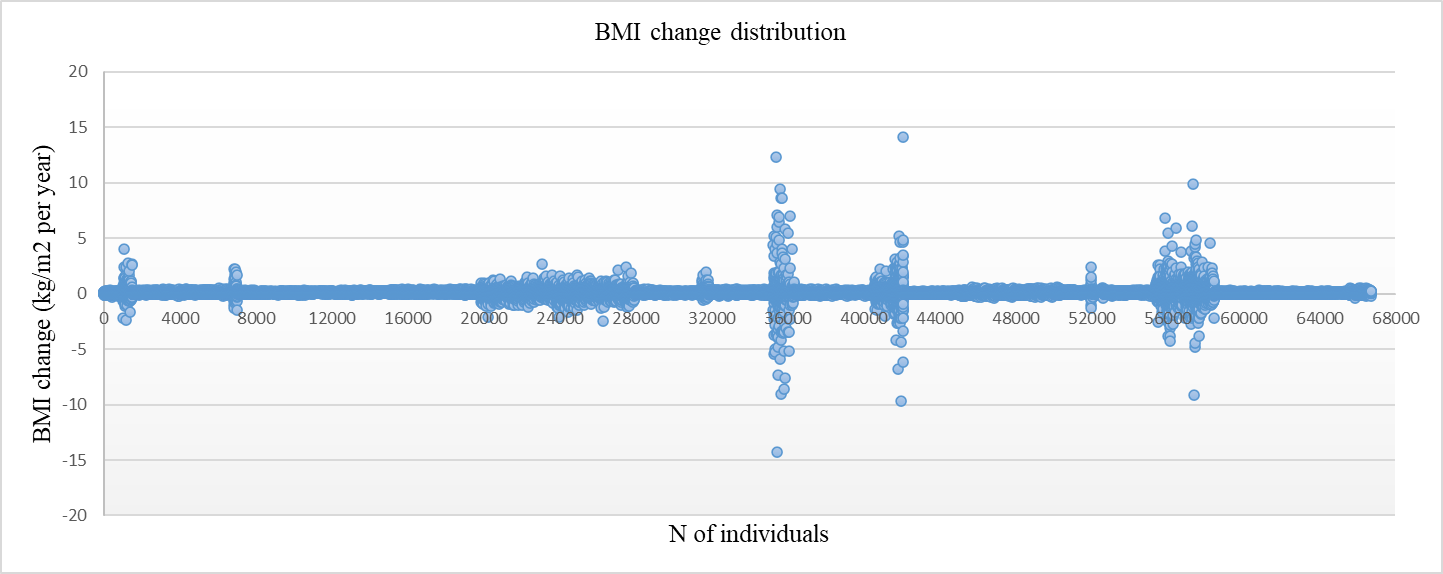
**Supplementary Figure 2**. BMI changes distribution for identifying outliers.

**Caption:** Scatter plot displaying BMI change values across individuals is shown in the figure. Individuals with BMI change values greater than 3 standard deviations from the mean were considered outliers and removed prior to analysis. **Abbreviations:** BMI: body mass index

**Supplementary Figure 3:** Flow chart illustrating the selection and inclusion process of participants in the current study.

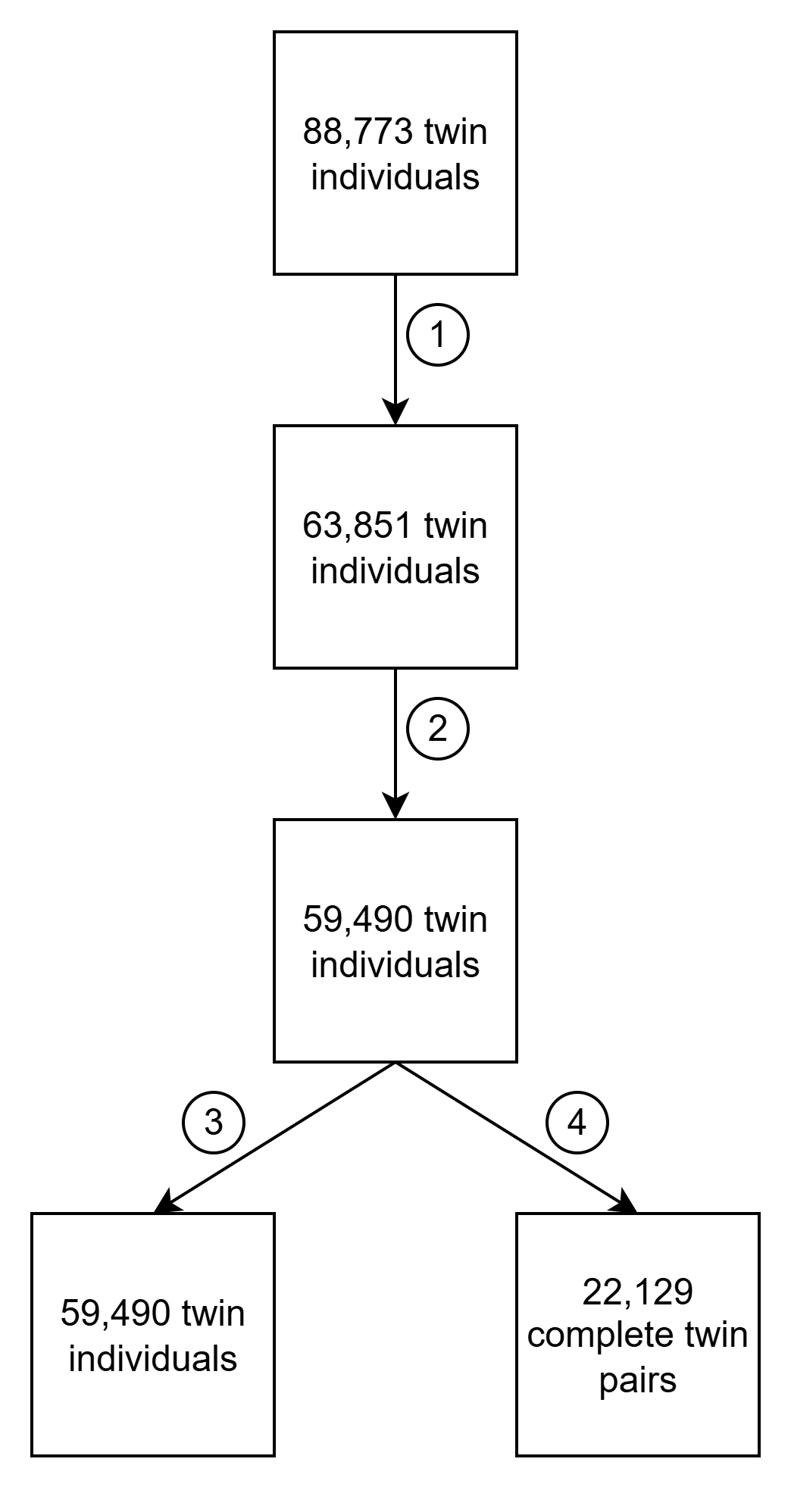

**Caption**: The study began with 88,730 individuals drawn from cohorts that included information on educational attainment and repeated BMI measurements. (1) Participants with educational data reported only before age 30 were excluded. (2) We then examined the distribution of baseline BMI and BMI change, excluding those with values exceeding three standard deviations from the mean. (3) Associations at the individual level were analyzed using linear mixed-effects (LME) models. (4) Finally, structural equation modelling—both univariate and bivariate—was conducted using complete twin pairs.

**Supplementary Table 1.** Associations of EA with baseline BMI (kg/m^2^) and BMI change (kg/m^2^ per ten years) in each of the cohort included in the study, by sex.

|  | **Men** | | | **Women** | | |
| --- | --- | --- | --- | --- | --- | --- |
|  | **β** | **95%CI** | | **β** | **95%CI** | |
|  |  | **LL** | **UL** |  | **LL** | **UL** |
| *Europe* |  |  |  |  |  |  |
| Finnish old cohort | N= 9,879 | | | N= 11,059 | | |
| Baseline BMI | -0.09 | -0.11 | -0.07 | -0.17 | -0.19 | -0.14 |
| BMI change | -1.72e-03 | -2.71e-03 | 1.69e-03 | -3.92e-03 | -4.05e-03 | 1.80e-03 |
| Italian Twin Registry | N= 422 | | | N= 858 | | |
| Baseline BMI | -0.12 | -0.27 | 0.03 | -0.17 | -0.30 | -0.05 |
| BMI change | 1.26e-03 | -0.01 | 0.01 | -0.08 | -0.17 | 0.01 |
| Murcia Twin register | N= 467 | | | N= 831 | | |
| Baseline BMI | -0.09 | -0.17 | -0.02 | -0.16 | -0.23 | -0.10 |
| BMI change | -0.10 | -0.20 | 8.59e-04 | -0.12 | -0.20 | -0.01 |
| Netherlands Twin Register | N= 1,418 | | | N= 2,542 | | |
| Baseline BMI | -0.01 | -0.05 | 0.04 | -0.02 | -0.06 | 0.04 |
| BMI change | -2.19e-03 | -4.54e-03 | 1.53e-04 | -1.07e-03 | -3.10e-03 | 9.51e-03 |
| Swedish Twin Register | N= 3,704 | | | N= 4,550 | | |
| Baseline BMI | -0.11 | -0.13 | -0.08 | -0.13 | -0.16 | -0.10 |
| BMI change | -2.04e-03 | -7.80e-03 | 0.01 | -0.02 | -0.03 | -0.01 |
| *North America* |  |  |  |  |  |  |
| Colorado Twin Registry | N= 649 | | |  | N= 829 |  |
| Baseline BMI | 0.11 | -0.12 | 0.34 | -0.10 | -0.21 | 1.55e-03 |
| BMI change | -0.01 | -0.07 | 0.08 | -0.02 | -0.03 | -0.01 |
| NAS-NRC Twin Registry | N= 8,692 | | | N= 0 | | |
| Baseline BMI | -0.01 | -0.02 | 0.01 | - | - | - |
| BMI change | -0.01 | -0.02 | -6.57e-03 | - | - | - |
| Vietnam Era Twin Study of Aging | N= 743 | | | N= 0 | | |
| Baseline BMI | -0.11 | -0.24 | 0.01 | - | - | - |
| BMI change | -0.04 | -0.10 | 0.07 | - | - | - |
| Washington State Twin Registry | N= 1,994 | | | N= 3,883 | | |
| Baseline BMI | -0.13 | -0.20 | -0.05 | -0.31 | -0.38 | -0.24 |
| BMI change | -0.01 | -0.08 | 0.05 | -0.03 | -0.10 | 0.03 |
| *Asia* |  |  |  |  |  |  |
| Korea Twin-Family Register | N= 77 | | | N= 123 | | |
| Baseline BMI | 0.02 | -0.05 | 0.10 | -0.09 | -0.16 | -0.02 |
| BMI change | 0.01 | -0.01 | 0.02 | -0.04 | -0.12 | 0.03 |
| *Australia* |  |  |  |  |  |  |
| Queensland Twin Register | N= 3,394 | | | N= 3,472 | | |
| Baseline BMI | -0.11 | -0.16 | -0.05 | -0.18 | -0.22 | -0.14 |
| BMI change | -0.06 | -0.10 | -0.03 | -0.05 | -0.08 | -0.01 |
| Crochan Q test | Cochran's Q | | p-value | Cochran's Q | | p-value |
| Baseline BMI | 120.97 | | 3.95e-20 | 125.38 | | 4.06e-22 |
| BMI change | 42.88 | | 2.35e-05 | 65.00 | | 4.04e-10 |

**Caption:** For the study of the associations, LME models were used. The models are: i) Baseline BMI~ Maximum Educational attainment + Zygosity + Baseline Age + (1|Family ID) and ii) BMI change~ Maximum Educational attainment + Zygosity + Baseline Age + Baseline BMI + (1|Family ID). The estimates are displayed along with the 95% confidence interval. Cochran’s Q tests were performed to assess heterogeneity of the associations across cohorts. **Abbreviations:** BMI: Body mass index; CI: confidence interval; LL: lower limit; UL: upper limit; NAS-NRC: National Academy of Sciences-National Research Council.

**Supplementary Table 2.** Sensitivity analysis for the associations of educational attainment (in standardized education years) with BMI baseline (kg/m^2^) and BMI change (kg/m^2^ per decade) in pooled data by sex.

|  | **Men (N=7,360)** | | | **Women (N=7,642)** | | |
| --- | --- | --- | --- | --- | --- | --- |
|  | **β** | **95%CI** | | **β** | **95%CI** | |
|  |  | **LL** | **UL** |  | **LL** | **UL** |
| Baseline BMI | -0.06 | -0.07 | -0.05 | -0.13 | -0.15 | -0.11 |
| BMI change | -0.01 | -0.02 | -4.25e-03 | -0.01 | -0.02 | -1.12e-03 |

**Caption:** Associations for the sensitivity analysis including smoking behaviour as a covariate were carried out using LME models. The models are: i) BMI baseline~ Maximum Educational attainment + cohort ID + baseline smoking status + Zygosity + Baseline Age + (1|Family ID) and ii) BMI change~ Maximum Educational attainment + cohort ID + baseline smoking status + Zygosity + Baseline Age + BMI baseline + (1|Family ID). The estimates are displayed along with the 95% confidence interval. **Abbreviations:** BMI: Body mass index; CI: confidence interval; LL: lower limit; UL: upper limi

**Supplementary Table 3.** Sensitivity analysis for the associations of educational attainment (in standardized education years) with BMI baseline (kg/m^2^) and BMI change (kg/m^2^ per decade) at different stages of the adulthood in pooled data by sex.

|  | **Men** | | | **Women** | | |
| --- | --- | --- | --- | --- | --- | --- |
|  | **β** | **95%CI** | | **β** | **95%CI** | |
|  |  | **LL** | **UL** |  | **LL** | **UL** |
| Middle to late adulthood (30-50 years old) | N = 9,049 | | | N = 10,402 | | |
| Baseline BMI | -0.09 | -0.11 | -0.07 | -0.12 | -0.15 | -0.10 |
| BMI change | -5.69e-03 | -0.01 | 4.44e-03 | -0.02 | -0.03 | -8.45e-03 |
| Middle age (51-65 years old) | N = 1,093 | | | N = 1,799 | | |
| Baseline BMI | -0.04 | -0.11 | 0.01 | -0.09 | -0.17 | -0.02 |
| BMI change | -0.04 | -0.10 | 0.01 | -0.10 | -0.17 | -0.03 |
| Old age (>66 years old) | N = 171 | | | N = 374 | | |
| Baseline BMI | -0.12 | -0.27 | 0.03 | -0.24 | -0.41 | -0.08 |
| BMI change | -0.11 | -0.25 | 0.02 | -0.02 | -0.17 | 0.13 |

**Caption:** Sensitivity analyses dividing adulthood into different stages were performed using linear mixed-effects (LME) models, specified identically to the main association analyses. Outcomes were baseline BMI or BMI change, with educational attainment as the primary covariate. Models were adjusted for zygosity, cohort ID and baseline age; baseline BMI was additionally included as a covariate in the model of BMI change. The estimates are displayed along with the 95% confidence interval. **Abbreviations:** BMI: Body mass index; CI: confidence interval; LL: lower limit; UL: upper limit.

**Supplementary Table 4.** Intraclass correlations for baseline body mass index (BMI), its changes (kg/m^2^ per ten years) and standardized education years in pooled data by sex and zygosity.

| Variables of study | Men | | | | | | Women | | | | | | Opposite sex dizygotic (N=1,729) | | |
| --- | --- | --- | --- | --- | --- | --- | --- | --- | --- | --- | --- | --- | --- | --- | --- |
|  | MZ  (N= 4,818) | | | DZ  (N=6,214) | | | MZ  (N=4,044) | | | DZ  (N=5,324) | | |  |  |  |
|  | Estimate | 95% CI | | Estimate | 95% CI | | Estimate | 95% CI | | Estimate | 95% CI | | Estimate | 95% CI | |
|  |  | LL | UL |  | LL | UL |  | LL | UL |  | LL | UL |  | LL | UL |
| BMI baseline | 0.78 | 0.77 | 0.79 | 0.57 | 0.55 | 0.59 | 0.77 | 0.76 | 0.78 | 0.55 | 0.53 | 0.57 | 0.37 | 0.32 | 0.41 |
| BMI change | 0.11 | 0.08 | 0.14 | 0.08 | 0.07 | 0.09 | 0.19 | 0.16 | 0.22 | 0.13 | 0.11 | 0.15 | 0.11 | 0.06 | 0.15 |
| Standardized education years | 0.70 | 0.69 | 0.71 | 0.52 | 0.50 | 0.54 | 0.72 | 0.71 | 0.74 | 0.48 | 0.46 | 0.50 | 0.36 | 0.32 | 0.40 |

**Caption**: Intraclass correlations for baseline BMI, BMI changes and standardized education years in pooled data from Europe, Asia, Australia, and North America are summarized by sex and zygosity with correlation coefficients besides their 95% confidence interval. All the correlations are highly significant (all p<0.005). Although only pooled ICCs are shown, MZ correlations were consistently less than twice DZ correlations supporting low heterogeneity and justifying pooling. **Abbreviations:** MZ: Monozygotic; DZ: Dizygotic; CI: Confidence interval; LL: Lower limit; UL: Upper limit.

**Supplementary Table 5.** Model fit statistics of baseline body mass index (BMI), its changes (kg/m^2^ per decade) and educational attainment in the pooled data comparing different genetic models.

| Variables of study | Full ACE model (reference model) | | ACE model without sex-specific genetic effect (1) | | ACE model with same parameter estimates for men and women (2) | | Full AE model (3) | | AE model without sex-specific genetic effect (4) | | AE model with same parameter estimates for men and women (3) | |
| --- | --- | --- | --- | --- | --- | --- | --- | --- | --- | --- | --- | --- |
|  | -2LL | d.f | Δ -2 LL | p value | Δ -2 LL | p value | Δ -2 LL | p value | Δ -2 LL | p value | Δ -2 LL | p value |
| Baseline BMI | 21186.33 | 4225 | 0.49 | 0.48 | 370.05 | 6.78e-48 | 0.38 | 0.53 | 62.46 | 2.71e-15 | 73.01 | 1.39e-16 |
| BMI change | 16800.42 | 4225 | 0.61 | 0.42 | 200.34 | 5.48e-26 | 0.29 | 0.37 | 100.24 | 8.08e-17 | 207.44 | 6.31e-29 |
| EA | 21010.21 | 4225 | 7.65 | 5.94e-03 | 89.59 | 2.68e-19 | 31.29 | 8.25e-12 | 53.75 | 2.14e-12 | 18.22 | 1.96e-05 |

**Caption:** Full ACE model is compared against the rest of the models displaying the -2 log likelihood and degrees of freedom for the reference model and the differences in -2log likelihood and the p value in the models compared with the reference one. **Abbreviations:** -2LL (-2 log-likelihood); d.f. (degrees of freedom); Δ (change); ACE (additive genetic/ shared environment/ unique environment) model; AE (additive genetic/ unique environment) model. (1) Compared to full ACE model (Δ d.f. 1); (2) Compared to the ACE model without sex-specific genetic effect (Δ d.f. 1); (3) Compared to the full ACE model (Δ d.f. 2); (4) Compared to the full AE model (Δ d.f. 1); (5) Compared to the AE model without sex-specific genetic effect (Δ d.f. 1)

**Supplementary Table 6**. Contribution of additive genetic and unique environmental factors to the previously identified associations between EA and BMI trajectory components (Baseline BMI and BMI change) at cohort specific level analysis, by sex.

| **Variables of study** | **Men** | | | | | | **Women** | | | | | |
| --- | --- | --- | --- | --- | --- | --- | --- | --- | --- | --- | --- | --- |
|  | **Additive genetic correlation** | | | **Unique environmental correlations** | | | **Additive genetic correlation** | | | **Unique environmental correlation** | | |
|  | **r_A_** | **95% CI** | | **r_E_** | **95% CI** | | **r_A_** | **95% CI** | | **r_E_** | **95% CI** | |
|  |  | **LL** | **UL** |  | **LL** | **UL** |  |  |  |  |  |  |
| *Europe* |  |  |  |  |  |  |  |  |  |  |  |  |
| Finnish old cohort |  |  |  |  |  |  |  |  |  |  |  |  |
| Baseline BMI | -0.16 | -0.19 | -0.13 | -0.04 | -0.10 | -0.01 | -0.20 | -0.22 | -0.18 | 0.03 | -0.02 | 0.08 |
| BMI change | - | - | - | - | - | - | - | - | - | - | - | - |
| Italian Twin Registry |  |  |  |  |  |  |  |  |  |  |  |  |
| Baseline BMI | - | - | - | - | - | - | -0.90 | -1.00 | -0.78 | -0.24 | -0.52 | 0.05 |
| BMI change | - | - | - | - | - | - | - | - | - | - | - | - |
| Murcia Twin register |  |  |  |  |  |  |  |  |  |  |  |  |
| Baseline BMI | -0.31 | -0.49 | -0.14 | 0.17 | -0.03 | 0.37 | -0.43 | -0.58 | -0.28 | 0.01 | -0.11 | 0.13 |
| BMI change | 0.08 | -0.87 | 0.87 | -0.01 | -0.23 | 0.20 | -0.12 | -0.44 | 0.10 | -0.01 | -0.13 | 0.10 |
| Netherlands Twin Register |  |  |  |  |  |  |  |  |  |  |  |  |
| Baseline BMI | 0.31 | 0.24 | 0.38 | 0.15 | 0.01 | 0.29 | 0.32 | 0.26 | 0.39 | 0.13 | -0.01 | 0.27 |
| BMI change | - | - | - | - | - | - | - | - | - | - | - | - |
| Swedish Twin Register |  |  |  |  |  |  |  |  |  |  |  |  |
| Baseline BMI | -0.25 | -0.29 | -0.21 | 0.03 | -0.02 | 0.08 | -0.25 | -0.28 | -0.22 | 0.03 | -0.01 | 0.07 |
| BMI change | - | - | - | - | - | - | 0.01 | -0.04 | 0.05 | -0.01 | -0.05 | 0.03 |
| *North America* |  |  |  |  |  |  |  |  |  |  |  |  |
| Colorado Twin Register |  |  |  |  |  |  |  |  |  |  |  |  |
| BMI baseline | - | - | - | - | - | - | - | - | - | - | - | - |
| BMI change | - | - | - | - | - | - | - | - | - | - | - | - |
| NAS-NRC Twin Registry |  |  |  |  |  |  |  |  |  |  |  |  |
| BMI baseline | - | - | - | - | - | - | - | - | - | - | - | - |
| BMI change | -0.16 | -0.21 | -0.12 | 0.01 | -0.02 | 0.05 | - | - | - | - | - | - |
| Vietnam Era Twin Study of Aging |  |  |  |  |  |  |  |  |  |  |  |  |
| BMI baseline | - | - | - | - | - | - | - | - | - | - | - | - |
| BMI change | - | - | - | - | - | - | - | - | - | - | - | - |
| Washington State Twin Registry |  |  |  |  |  |  |  |  |  |  |  |  |
| Baseline BMI | -0.09 | -0.20 | 0.02 | -0.13 | -0.20 | -0.03 | -0.04 | -0.15 | 0.09 | -0.16 | -0.23 | -0.05 |
| BMI change | - | - | - | - | - | - | - | - | - | - | - | - |
| *Asia* |  |  |  |  |  |  |  |  |  |  |  |  |
| Korea Twin-Family Register |  |  |  |  |  |  |  |  |  |  |  |  |
| Baseline BMI | - | - | - | - | - | - | -0.12 | -0.30 | 0.07 | -0.27 | -0.45 | -0.02 |
| BMI change | - | - | - | - | - | - | - | - | - | - | - | - |
| *Australia* |  |  |  |  |  |  |  |  |  |  |  |  |
| Queensland Twin Register |  |  |  |  |  |  |  |  |  |  |  |  |
| Baseline BMI | -0.17 | -0.28 | -0.05 | -0.01 | -0.09 | 0.08 | -0.12 | -0.21 | -0.04 | -0.10 | -0.15 | -0.04 |
| BMI change | -0.19 | -0.31 | -0.07 | -0.02 | -0.11 | 0.06 | -0.24 | -0.39 | -0.12 | 0.01 | -0.04 | 0.06 |
| Crochan Q test | Cochran's Q | | p-value | Cochran's Q | | p-value | Cochran's Q | | p-value | Cochran's Q | | p-value |
| Baseline BMI | 195.14 | | 2.05e-39 | 21.27 | | 3.38e-03 | 441.93 | | 1.52e-89 | 46.10 | | 1.37e-06 |
| BMI change | 1.76 | | 0.62 | 14.72 | | 2.06e-03 | 12.59 | | 5.58e-03 | 0.49 | | 0.92 |

**Caption**: Additive genetic (r_A_) and unique environmental (r_E_) correlations of EA with Baseline BMI and BMI change were obtained for the previously found significant associations at cohort specific level using the genetic twin models, which decompose phenotypic covariation into genetic and environmental effects. Cochran’s Q tests were performed to assess heterogeneity of these correlations across cohorts and sex. **Abbreviations**: r_A_: Additive genetic correlation coefficient; r_E_; Specific environmental correlation coefficient; LL: Lower limit; UL: Upper limit.
